# Japanese Questionnaire Survey Examining the Invasiveness of Phlebotomy in Medical Research

**DOI:** 10.1101/2025.04.09.25325435

**Authors:** Yusuke Ebana, Megumu Yokono, Masayuki Yoshida

## Abstract

**Introduction:** Phlebotomy, or venipuncture, is a minimally invasive procedure commonly used in routine medical practice and research for blood collection. While generally safe when conducted according to standardized measures, it carries risks like hematomas and vasovagal reflex for patients, and bacterial infection for healthcare workers. The invasiveness of phlebotomy depends on factors like patient age, volume of blood drawn, and collection frequency. Ethical guidelines, including those in Japan, require informed consent and safety measures in research, emphasizing the need for scientific or societal justification. This study aims to assess phlebotomy’s invasiveness across various research scenarios to better understand its implications.

**Methods:** An online questionnaire survey was conducted among Certified Research Ethics Committee Professionals (CRePs) in Japan, who are experts in research ethics. These professionals check research protocols to ensure compliance with legal requirements and standards and provide necessary information for ethical committee reviews. The survey was conducted in September 2024. The questionnaire included four scenarios: healthy adult, healthy minor, adult patient, and minor patient, asking respondents to define the invasiveness threshold based on the amount of blood drawn or the number of collections.

**Result:** A total of 77 CRePs participated in the survey, with most respondents serving on the ethics committees of academic institutions (72%) and possessing significant experience, 73% reported serving 5 or more years. For healthy adults, 50% of respondents considered 20 mL of blood invasive, while 30% identified 50 mL as the threshold; for adult patients, the responses were similar, though more considered 200 mL or always minimally invasive. Regarding frequency, a few times was the most common threshold for invasiveness, followed by once and several times, showing higher tolerance for patients. For minors, responses leaned more toward always invasive, especially for healthy children, with thresholds increasing for patient cases. Age-related invasiveness criteria varied widely, with significant caution for non-patients.

**Discussion:** This study explored the invasiveness of phlebotomy in medical research under Japanese ethical guidelines, highlighting its classification as a minimally invasive procedure. Phlebotomy’s invasiveness is influenced by factors such as patient age, volume of blood drawn, and number of collections, with ethical considerations being particularly important in research contexts. The findings revealed differing opinions among CRePs, who are well-versed in research protocols, especially regarding minor versus adult cases. Interventions for mitigating pain and anxiety during phlebotomy, particularly in children, were emphasized, along with the need to distinguish between research-related phlebotomy and blood donation standards due to differing objectives.

**Conclusion:** Opinions regarding the invasiveness of phlebotomy for research purposes vary by the amount of blood drawn and number of collections. In general, phlebotomy is considered invasive if the blood is drawn from a minor.

## Introduction

Phlebotomy involves inserting a needle into a vein (venipuncture) to draw blood.[1] Patients may consider the pain and discomfort caused by venipuncture a source of anxiety and other forms of stress. Venipuncture exposes the patients to the risk of adverse events such as hematoma and vasovagal reflex, as well. For healthcare workers, venipuncture poses the risk of bacterial infection.[3] In regular medical settings, phlebotomy is conducted safely; institutions conduct phlebotomy safely by following standardized measures based on an established methodology. Since blood tests form an essential part of routine medical practice, collecting blood samples for research purposes is typically considered an extension of a doctor’s medical practice. From the Ethical, Legal and Social Issues (ELSI) perspective, we can observe the difference that “voluntary” is required for blood collection in medical practice, while “self-initiation” is required for blood collection in research. Currently, guidelines of medical research define puncture on human subjects an invasive act. However, phlebotomy is generally classified as a minimally invasive procedure. Few phlebotomy encounters are unsuccessful: In one study, 93.2% of inpatient phlebotomy encounters were successful.[3] However, due to the preponderance of people who dread phlebotomy encounters, it is necessary to consider the perspectives of parties other than phlebotomists.

Medical researchers have an ethical duty to protect research subjects, as enshrined in the Declaration of Helsinki, as well as national legislation and ethical guidelines.[4][5][6] Phlebotomy is a common procedure in medical research. Healthcare workers collect blood, as part of their routine practice, to perform blood tests (e.g., blood cell, biochemical, and immunologic tests) that help determine whether the patient has a disease or measure the severity of disease. Medical researchers collect blood for various purposes such as evaluating the pharmacokinetic properties or efficacy of a tested drug, tracking a disease-related biomarker, or analyzing correlation with disease severity.

The glossary section of Japan’s Ethical Guidelines for Medical and Health Research Involving Human Subjects provides a definition of the term “invasiveness”[5] and some examples of invasive” procedures, including puncture, incision, administering drugs, irradiation, and presenting traumatic images or text. In medical research, it is necessary to identify the procedures that are invasive because they require the subject’s informed consent and entail the duty to ensure the subject’s safety. Further, they require the researcher to justify the act on the grounds of its societal benefits or scientific rationale.

By this standard, phlebotomy is generally considered minimally invasive because the risks associated with the safe conduction of phlebotomy in routine medical practice are minimal. However, it is wrong to assume that phlebotomy’s invasiveness is minimal in all cases. The invasiveness of a particular phlebotomy encounter will depend on the patient’s age, the amount of blood drawn, the number of blood collections, and situational variables.

Therefore, the purpose of the current study is to clarify the invasiveness of phlebotomy, particularly that conducted pursuant to a research protocol submitted to an ethical committee and institutional review board for approval, in medical research. To this end, multiple scenarios of such phlebotomy were envisaged and the invasiveness of each scenario was assessed.

## Method

### Participants

An online questionnaire survey was conducted among Certified Research Ethics Committee Professionals (CRePs), who are professionals of medical ethics in Japan. They serve on the ethical committees of academic, medical, or corporate institutions or manage the secretariats of institutional review boards.[7] In this role, they subject research proposals to legal requirements and standards and gather the information necessary for their ethical committee to review whether the research proposal should be approved.

In this study, the CRePs were notified about the questionnaire survey by email. The survey was conducted in September 2024, one month after notifying the professionals.

### Questionnaire on Phlebotomy

The questionnaire presented respondents with multiple phlebotomy scenarios: healthy adult, healthy minor, adult patient, and minor patient. In each case, the respondents were asked to describe the invasiveness threshold in terms of the amount of blood drawn or number of collections:

1. For research purposes, a research team wants to draw blood from a healthy adult. For a single phlebotomy encounter, how much blood will have to be drawn for you to consider the encounter invasive (as opposed to minimally invasive)?
2. For research purposes, a research team wants to draw blood from an adult patient who has regularly undergone phlebotomy for medical purposes. For a single phlebotomy encounter, how much blood will have to be drawn for you to consider the encounter invasive?
3. For research purposes, a research team wants to draw blood from a healthy adult. How many times will blood have to be drawn within a 24-hour period for you to consider the collections invasive? Reference: A patient with endocrine disease has three to six phlebotomy encounters.
4. For research purposes, a research team wants to draw blood from an adult patient who has regularly undergone phlebotomy for medical purposes. How many times will blood have to be drawn within a 24-hour period for you to consider the collections invasive?
5. For research purposes, a research team wants to draw blood from a healthy minor. For a single phlebotomy encounter, how much blood will have to be drawn for you to consider the encounter invasive?
6. For research purposes, a research team wants to draw blood from a non-adult patient. For a single phlebotomy encounter, how much blood will have to be drawn for you to consider the encounter invasive?
7. For research purposes, a research team wants to draw blood from a healthy minor. How young should the person be for you to consider the encounter invasive, regardless of the amount of blood drawn?
8. For research purposes, a research team wants to draw blood from a non-adult patient. How young should the person be for you to consider the phlebotomy encounter invasive, regardless of the amount of blood drawn?
9. Please write down any opinion you wish to share regarding phlebotomy for research purposes (you can leave this question blank if you wish).

## Results

### Respondent Details

A total of 77 CRePs responded to the survey. The majority of the respondents served on the ethics committees of academic institutions and, thereby, had many years of experience: For instance, 72% of the respondents served on the ethics committees of universities or other academic institutions, 17% the ethics committee of a hospital, and 10% the ethics committee of a for-profit corporation (see Figure 1). On being asked about their experience serving on an ethics committee, 49%, 24%, and 22% answered 5–10, 10 or more, and 3–5 years, respectively (see Figure 2).

**Figure 1.**
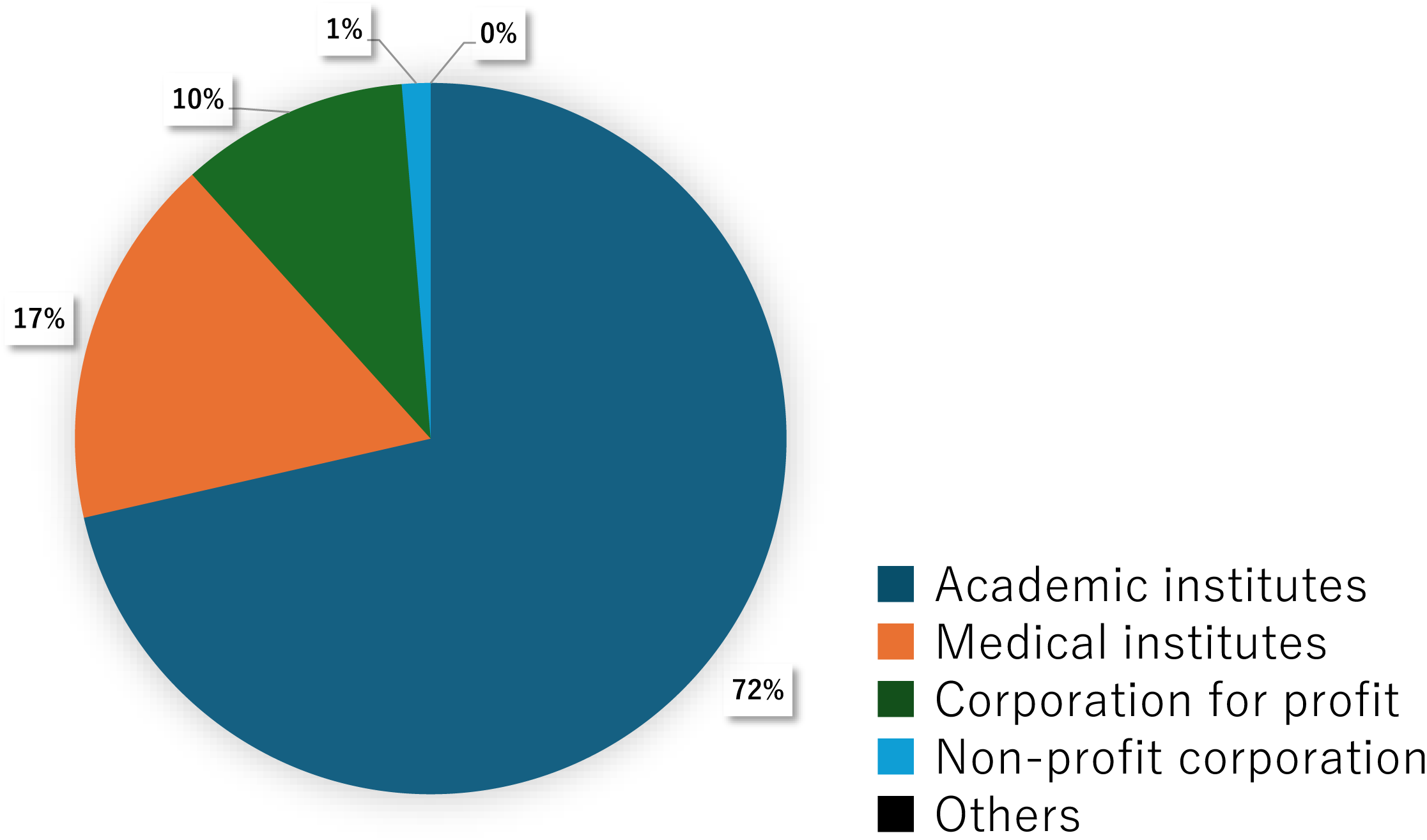
Organizational affiliations of Certified Research Ethics Committee Professionals (CReP) respondents

**Figure 2.**
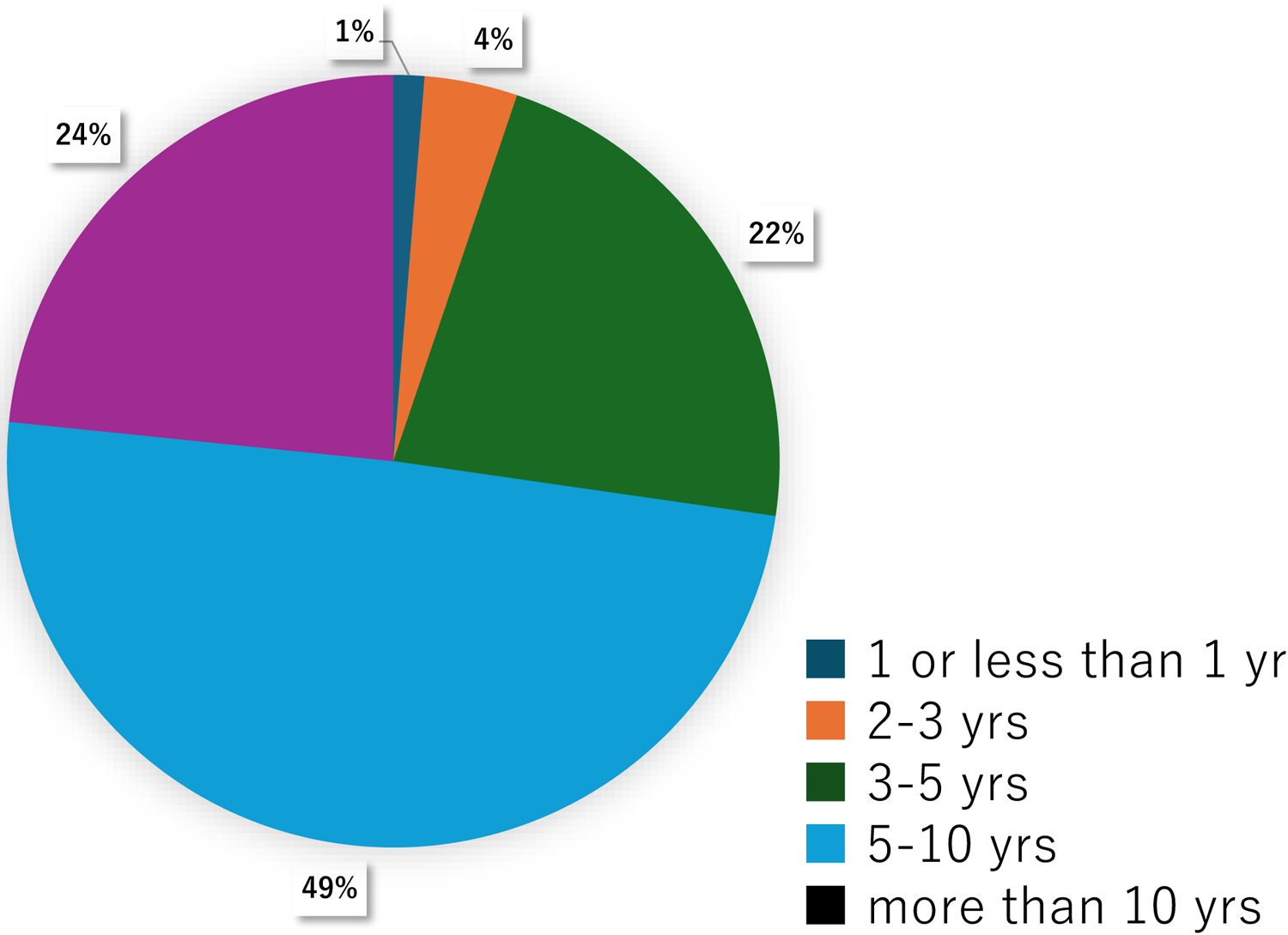
Years of experience of Certified Research Ethics Committee Professionals (CReP) respondents

### Research Phlebotomy in the Case of an Adult

A total of 77 responses to the invasiveness thresholds (the amount of blood, number of collections) in an adult were obtained. The current study differentiated between the case of an adult and that of a minor in light of an earlier report highlighting treatment discrepancies between adults and minors. Moreover, it differentiated the case of a patient and that of a healthy person on the assumption that CRePs’ opinions might vary between the cases.

When asked about a healthy adult, 50% of the respondents opined that a phlebotomy encounter would be invasive if it involved the drawing of at least 20 mL of blood, whereas 30% considered the drawing of at least 50 mL to be invasive (see Figure 3A). For an adult patient, “at least 20 mL” had a similarly high rate, at 49%. Further, “at least 50 mL” had, at 26%, a slightly lower rate than the healthy adult case, whereas “at least 200 mL” and “always minimally invasive (regardless of the amount drawn)” constituted a larger share compared to the previous case (see Figure 3B).

**Figure 3.**
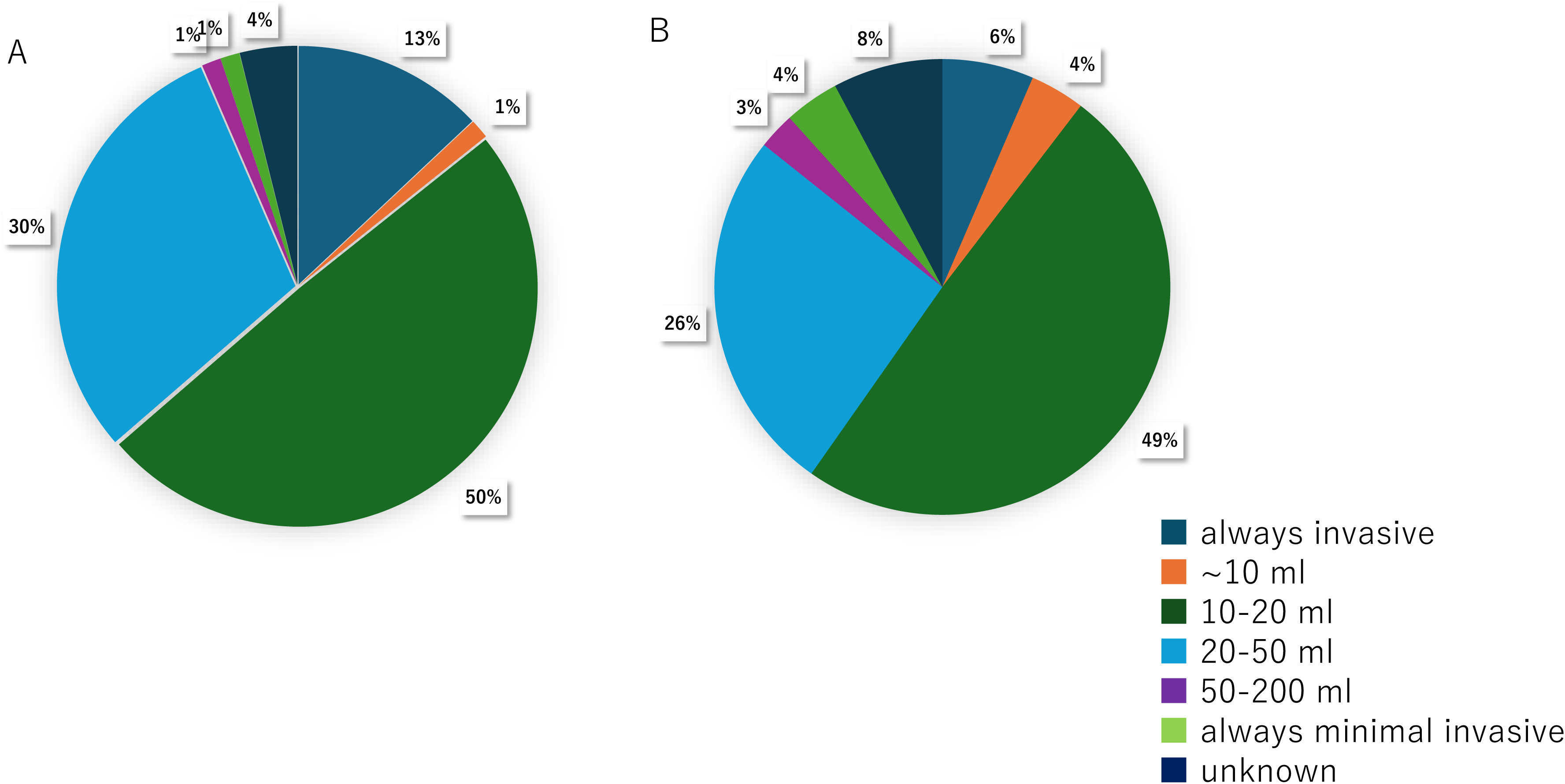
Distribution of Certified Research Ethics Committee Professionals’ responses regarding the threshold for considering phlebotomy for research purposes to be invasive; adult, amount of blood drawn: (3A) Healthy adult, (3B) adult patient

For the number of blood collections, as well, respondents were asked to consider the scenario of a healthy adult and that of an adult patient separately. In medical checkups pursuant to Japan’s National Health Insurance Act, blood is generally drawn only once. However, sometimes, blood is drawn multiple times to diagnose patients with endocrinal or metabolic diseases. When asked about the number of times blood would have to be drawn for the collections to be considered invasive, 34% of respondents answered “2 or 3 times” (see Figure 4A). The second and third most popular answers were “once” (22%) and “4–6 times” (18%), respectively. For the case of an adult patient, the most popular answer was “2–3 times” (38%), followed by “4–6 times” (22%). In this case, 12% answered “once.” These results highlight CRePs’ belief that patients have greater phlebotomy tolerance than non-patients (see Figure 4B).

**Figure 4.**
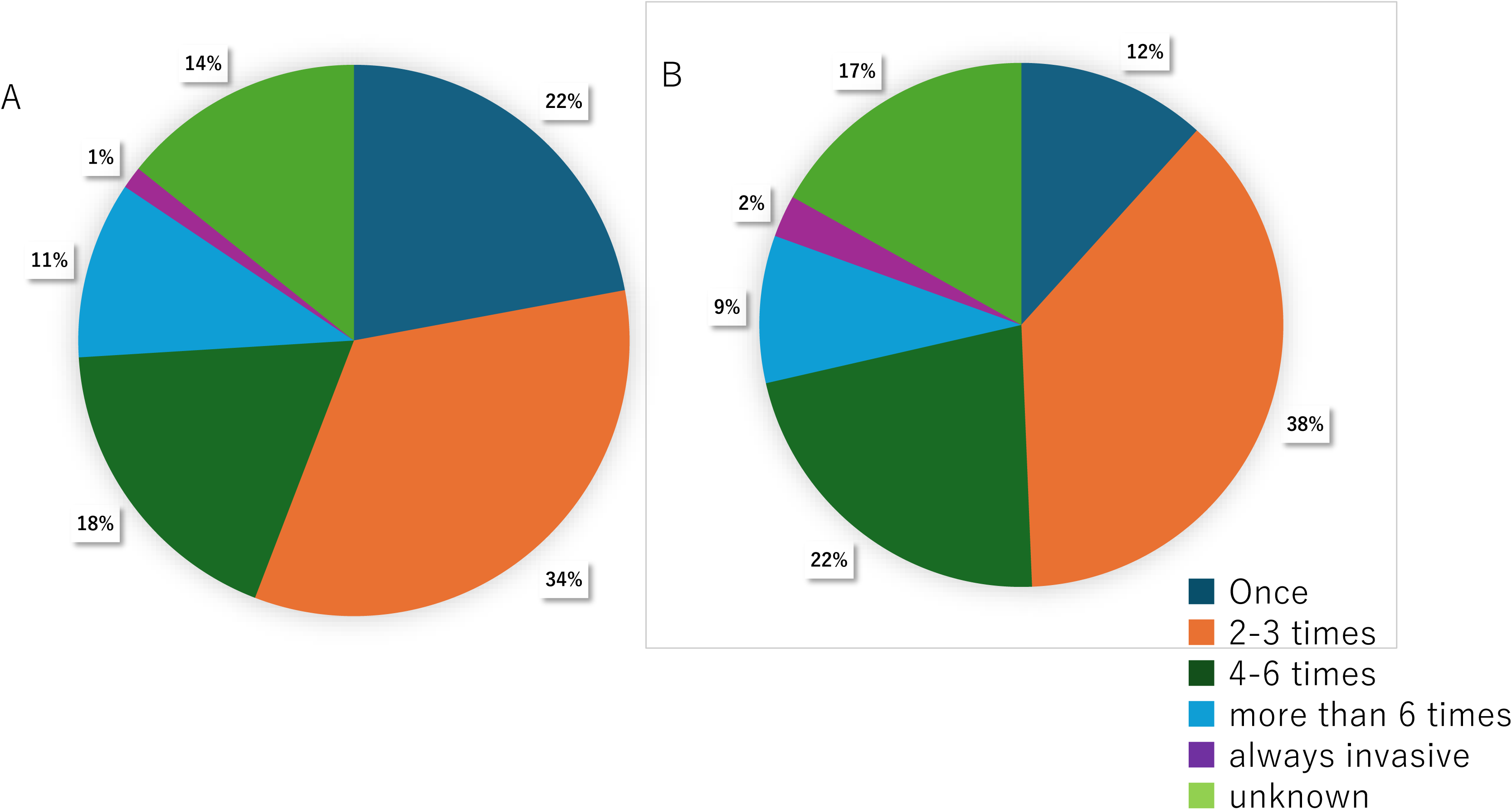
Distribution of Certified Research Ethics Committee Professionals’ responses regarding the threshold for considering phlebotomy for research purposes to be invasive; adult, number of collections: (4A) Healthy adult, (4B) adult patient

### Invasiveness of Research Phlebotomy in the Case of a Minor

For cases when the person was a minor, the questionnaire differentiated between the scenario where the person was a patient and that where the person was healthy. In both the scenarios, respondents were more likely to say “always invasive” compared to the adult scenarios. For a healthy child, “always invasive (regardless of the amount drawn)” and “at least 10 mL” each represented 26% of the responses. Further, only 4% of the respondents answered “always minimally invasive (regardless of the amount drawn),” which was a lower share than in the adult scenarios. Further, the respondents indicated higher blood volume thresholds for a child patient than they did for a healthy child (see Figures 5A and 5B).

**Figure 5.**
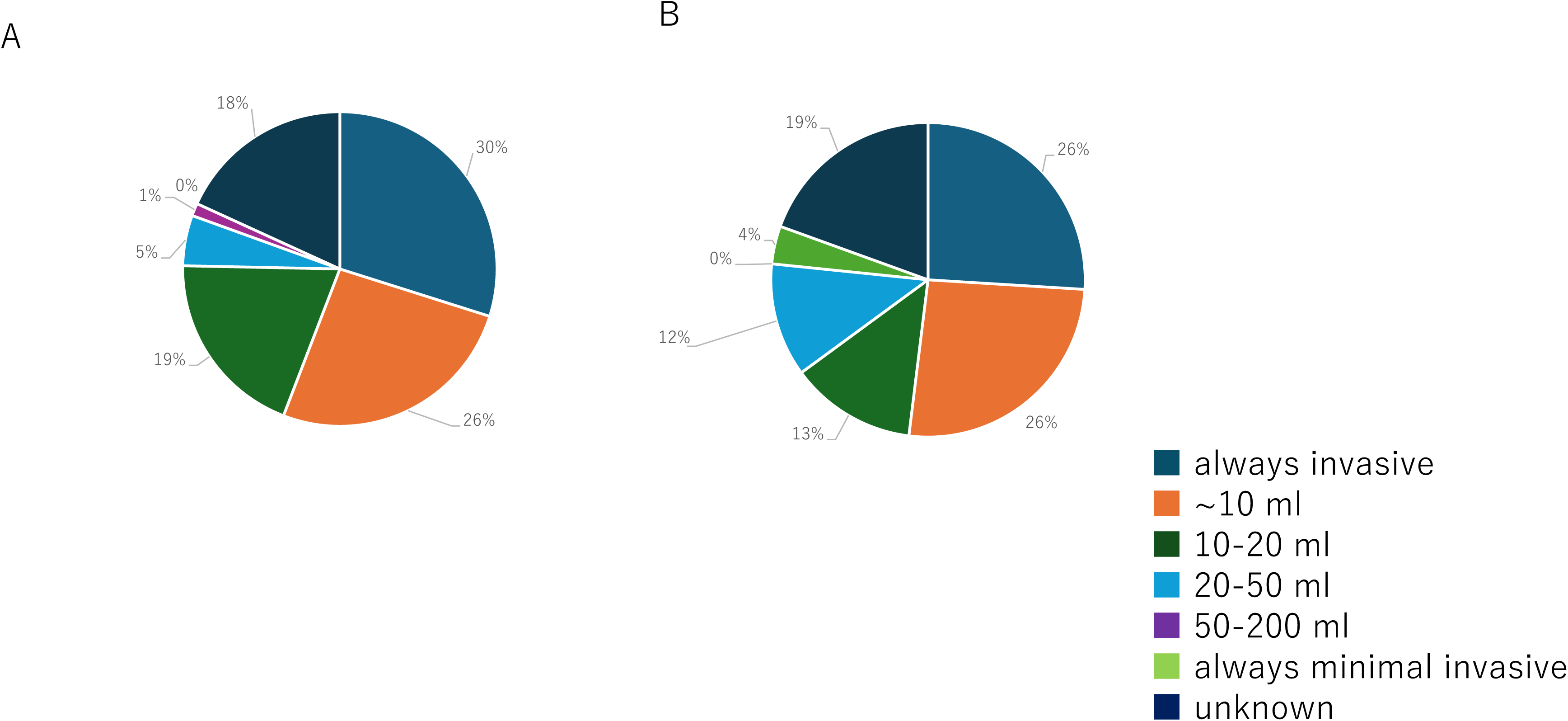
Distribution of Certified Research Ethics Committee Professionals’ responses regarding the threshold for considering phlebotomy for research purposes to be invasive; minor, amount of blood drawn: (5A) Healthy minor, (5B) non-adult patient

Finally, on being asked to state the age at which phlebotomy was invasive, the respondents likely answered “always invasive (regardless of the minor’s age)” when the child was a non-patient. The responses varied widely in age threshold for determining invasiveness (see Figures 6A and 6B).

**Figure 6.**
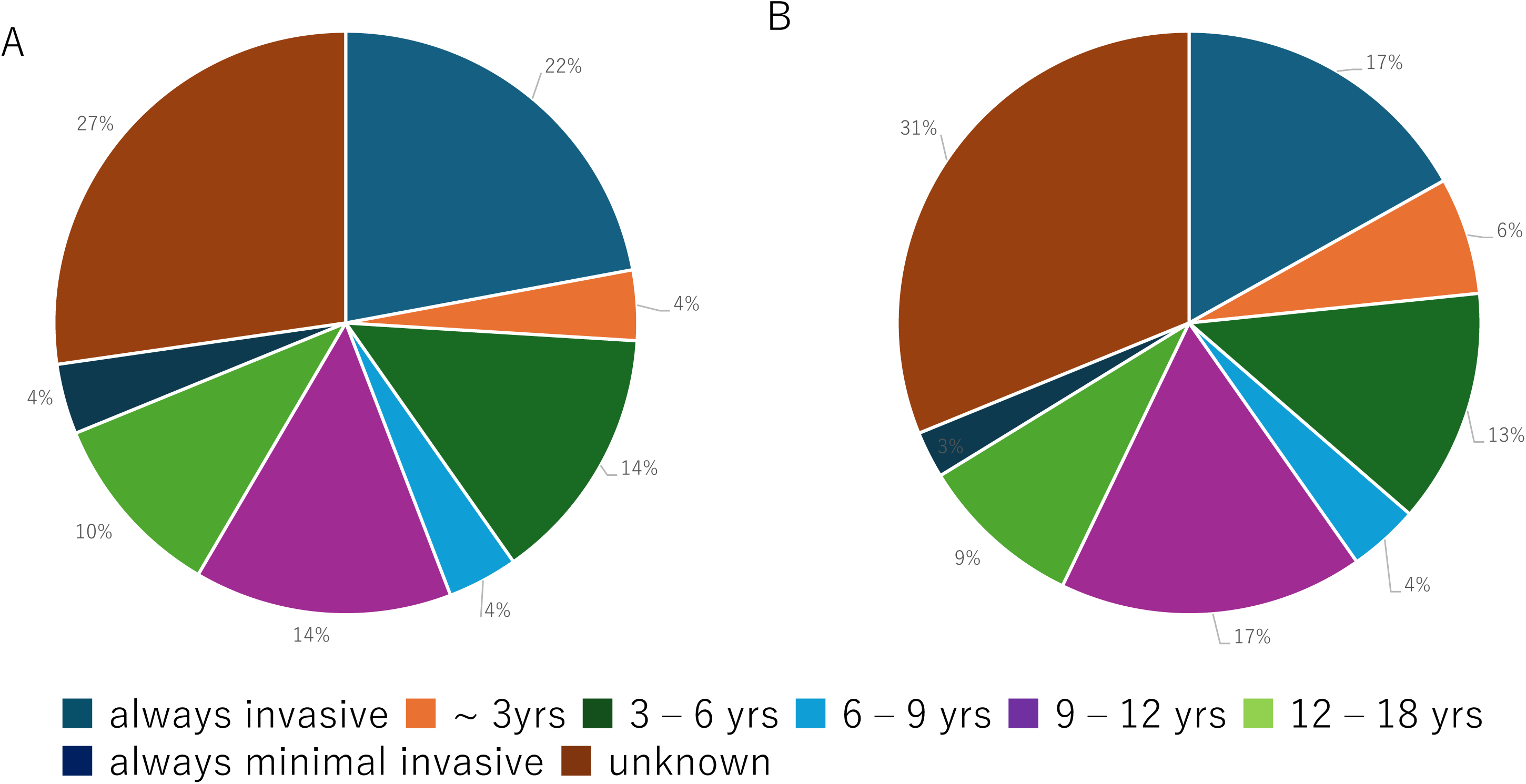
Distribution of Certified Research Ethics Committee Professionals’ responses regarding the threshold for considering phlebotomy for research purposes to be invasive; minor, age: (6A) Healthy minor, (6B) non-adult patient

## Discussion

This study examined the invasiveness of drawing blood in medical research, which is conducted pursuant to Japanese ethical guidelines. Blood tests form an essential part of routine medical practice. Therefore, collecting blood samples for research purposes is typically considered an extension of a doctor’s medical practice. Whereas institutional guidelines define the conduction of venipuncture on human subjects an invasive act, phlebotomy is generally classified as a minimally invasive procedure. Only few phlebotomy encounters are unsuccessful: According to one study, 93.2% of inpatient phlebotomy encounters are successful. However, due to the preponderance of people who dread phlebotomy encounters, it is necessary to consider the perspectives of those other than phlebotomists. Hence, this study was conducted.

As professionals engaged in ethical review, CRePs play a back-office, rather than a front-line medical, role. Indeed, many CRePs have no medical license.[7] Although their opinions on phlebotomy are similar to those of patients and research participants, they have a better understanding of research protocols than the latter.

The current study’s results revealed the differing opinions among CReP respondents regarding invasiveness thresholds in terms of the amount of blood drawn and number of collections. The respondents were more likely to consider phlebotomy invasive in the case of a minor than an adult. The number of interventions has been suggested for mitigating the pain and anxiety experienced by children during medical phlebotomy.[8][9][10] For adults, the literature is scarcer; however, reports suggest that childhood medical experiences can cause anxiety about medical situations in adulthood.[11][12]

Regarding phlebotomy for children, the determination of invasiveness may depend on age and weight, as well as the amount drawn and number of collections. Hence, decisions on invasiveness are shaped by many factors.

The Japanese Red Cross states that 200, 400, or 600 mL of blood should be drawn during a donation depending on criteria such as age, weight, and blood pressure.[13] In phlebotomy for research purposes, few consider it acceptable to draw 200 mL of blood or more. A consideration of the invasiveness of phlebotomy for research purposes indicates that the application of invasiveness standards in blood donations (in which blood is taken to save the lives of others) and medical phlebotomy is probably unwise, since the objectives in these cases are different from the aims of research.

Regarding the number of collections, the current study revealed that phlebotomy was more likely to be considered invasive if it involved a high number of collections. Some respondents believed that a high number of collections would be minimally invasive if they were intended for medical purposes but invasive if they were for research needs. We observed the tendency that people in Japan have a wider tolerance for the frequency of phlebotomy and a stricter impression of blood quantity as for the purpose of research, compared with the rule of United States.

## Conclusions

Opinions regarding the invasiveness of phlebotomy for research purposes vary by the amount of blood drawn and number of collections. In general, phlebotomy is considered invasive if the blood is drawn from a minor.

## Limitations

The questionnaire survey was conducted on CRePs alone. Many of them serve on the ethics committees of universities or other academic institutions; accordingly, they have more encounters with the research protocols submitted by medical research teams than most individuals. Therefore, CRePs’ views may not reflect the general population’s perceptions.

## Data Availability

All data produced in the present study are available upon reasonable request to the authors

## Appendix

### Questionnaire Items

Question 1. For research purposes, a research team wants to draw blood from a healthy adult. For a single phlebotomy encounter, how much blood will have to be drawn for you to consider the encounter invasive (as opposed to minimally invasive)?

1. I will consider it invasive (as opposed to minimally invasive) regardless of the amount of blood drawn.
2. I will consider it invasive if at least 10 mL of blood is drawn.
3. I will consider it invasive if at least 20 mL of blood is drawn.
4. I will consider it invasive if at least 50 mL of blood is drawn.
5. I will consider it invasive if at least 200 mL of blood is drawn.
6. I will consider it minimally invasive regardless of the amount of blood drawn.
7. I’m not sure.

Question 2. For research purposes, a research team wants to draw blood from an adult patient who has regularly undergone phlebotomy for medical purposes. For a single phlebotomy encounter, how much blood will have to be drawn for you to consider the encounter invasive?

1. I will consider it invasive (as opposed to minimally invasive) regardless of the amount of blood drawn.
2. I will consider it invasive if at least 10 mL of blood is drawn.
3. I will consider it invasive if at least 20 mL of blood is drawn.
4. I will consider it invasive if at least 50 mL of blood is drawn.
5. I will consider it invasive if at least 200 mL of blood is drawn.
6. I will consider it minimally invasive regardless of the amount of blood drawn.
7. I’m not sure.

Question 3. For research purposes, a research team wants to draw blood from a healthy adult. How many times will blood have to be drawn within a 24-hour period for you to consider the collections invasive?

For your reference, a patient with endocrine disease has three to six phlebotomy encounters.

1. Once.
2. 2–3 times.
3. 4–6 times.
4. At least 6 times.
5. I will consider it minimally invasive regardless of the amount of blood drawn.
6. I’m not sure.

Question 4. For research purposes, a research team wants to draw blood from an adult patient who has regularly undergone phlebotomy for medical purposes. How many times will blood have to be drawn within a 24-hour period for you to consider the collections invasive?

1. Once.
2. 2–3 times.
3. 4–6 times.
4. At least 6 times.
5. I will consider it minimally invasive regardless of the amount of blood drawn.
6. I’m not sure.

Question 5. For research purposes, a research team wants to draw blood from a healthy minor. For a single phlebotomy encounter, how much blood will have to be drawn for you to consider the encounter invasive?

1. I will consider it invasive if at least 10 mL of blood is drawn.
2. I will consider it invasive if at least 20 mL of blood is drawn.
3. I will consider it invasive if at least 50 mL of blood is drawn.
4. I will consider it invasive if at least 200 mL of blood is drawn.
5. I will consider it invasive (as opposed to minimally invasive) regardless of the amount of blood drawn.
6. I’m not sure.

Question 6. For research purposes, a research team wants to draw blood from a non-adult patient. For a single phlebotomy encounter, how much blood will have to be drawn for you to consider the encounter invasive?

1. I will consider it invasive regardless of the amount of blood drawn.
2. I will consider it invasive if at least 10 mL of blood is drawn.
3. I will consider it invasive if at least 20 mL of blood is drawn.
4. I will consider it invasive if at least 50 mL of blood is drawn.
5. I will consider it minimally invasive regardless of the amount of blood drawn.
6. I’m not sure.

Question 7. For research purposes, a research team wants to draw blood from a healthy minor. How young will the person have to be for you to consider the phlebotomy encounter invasive, regardless of how much blood is drawn?

1. I will consider it invasive (as opposed to minimally invasive) regardless of the child’s age.
2. I will consider it invasive if the child is aged 3 or younger.
3. I will consider it invasive if the child is aged 6 or younger (pre-elementary age).
4. I will consider it invasive if the child is aged 9 or younger (lower-elementary age).
5. I will consider it invasive if the child is aged 12 or younger (elementary age).
6. I will consider it invasive if the child is aged 18 or younger (high-school age).
7. I will never consider it invasive (as opposed to minimally invasive) regardless of the child’s age.
8. I’m not sure.

Question 8. For research purposes, a research team wants to draw blood from a non-adult patient. How young will the person have to be for you to consider the phlebotomy encounter invasive, regardless of how much blood is drawn?

1. I will consider it invasive (as opposed to minimally invasive) regardless of the child’s age.
2. I will consider it invasive if the child is aged 3 or younger.
3. I will consider it invasive if the child is aged 6 or younger (pre-elementary age).
4. I will consider it invasive if the child is aged 9 or younger (lower-elementary age).
5. I will consider it invasive if the child is aged 12 or younger (elementary age).
6. I will consider it invasive if the child is aged 18 or younger (high-school age).
7. I will never consider it invasive (as opposed to minimally invasive) regardless of the child’s age.
8. I’m not sure.

